# By the Numbers: Results from A Quantitative Study of Health Information User Experience in People with and Without Disabilities

**DOI:** 10.1101/2025.10.01.25337080

**Authors:** Sonal Sathe, Nathaniel Porter

## Abstract

People with and without disabilities sought information online during the initial onset of COVID-19 during the official public health emergency from March 2020 to May 2023. After the public health emergency ended, though, there has been little information to assess the user experience of online health information seeking with respect to COVID-19, which remains a public health concern, in people with and without disabilities. The goal of this research is to assess the extent to which online content about COVID-19 is both usable and satisfying to people with disabilities and compare this to people without disabilities. A cross-sectional survey was used for this purpose. Results indicate that people with disabilities are more likely to find online content either unusable or dissatisfying compared to people without disabilities. Further work should recruit from a more racially and socio-economically diverse participant pool. Further work should also involve qualitative research with technology users with disabilities assessing specific challenges they may encounter.

## Introduction

The social model of disability calls for the elimination of barriers to participation and inclusion. The focus for this work is not on a specific type of disability, but disability as a phenomenon more generally and as a category of understanding and identity. Disability as a category of identity and understanding has been recognized by various research scholars as an important part of identity development. Disability studies scholars like Margaret Murugami have long asserted that disability as an identity helps a sense of individuality develop apart from an idea of disability as tragedy (2009). Recently, researchers such as Anjali Forber-Pratt and colleagues (2020) have also conducted their own quantitative studies showing how disability identity development occurs and how disability identity can be useful as a category for both statistical analysis and for development of one’s sense of self and belonging. Both scholars kept the category and identity of disability broad. As such, this article will do the same.

This article seeks to understand the experiences of usability (or unusability) of certain technologies for people with disabilities and compare them to people without disabilities. We specifically seek to understand how people with disabilities may experience barriers to usability of technology. For this article, technology means web-based search engines and/or webpages that people with and without disabilities use to access information about COVID-19; these webpages and search engines may have AI-based tools embedded in them whether the user wants to use them or not. We question the idea that people with disabilities do not have any different experiences using technology compared to their counterparts without disabilities and use a cross-sectional survey and statistical analyses to highlight these differences.

### Health Information and its Intersection with Disability, User Experience Research, Universal Design, and Artificial Intelligence

Health information, defined as information intended for potential users of various health services (Demeris, 2016), can help people make decisions about preventive healthcare upon being accessed. Some of those decisions include vaccination against diseases like COVID-19. Since people with disabilities are at increased risk for severe illness and death from COVID-19 (Kendall et al, 2020), it is crucial for them to be able to access usable and accurate health information.

This health information can come through a variety of conduits, such as mobile phones and internet search engines (Cross, 2017). Internet search engines, via both browsers and direct search interfaces, are embedded in mobile phones and other devices, and these search engines may integrate AI-based tools. People use these engines to search for health information (Narayanan et al, 2025). Computer scientists Stuart Russell & Peter Norvig define AI as a field of research in computer science that develops and studies methods and software that enables machines to perceive their environment and use learning and intelligence to take actions that maximize their chances of achieving defined goals (2021). AI-generated content and online information may or may not be usable or even accurate, and there is a danger to human health and life when people begin trusting inaccurate AI generated health information. As an example, believing that lung reduction surgery might cure emphysema when it cannot do this could harm an already sick person (Shekar et al, 2025). Information being incorrect or unhelpful about something like COVID-19 may put health and lives at risk, especially for people with disabilities (Deal et al, 2023).

The landscape of health information in the 21^st^ century also means that health information, AI-generated or not, intersects disability, user experience research (UXR), and universal design (UD). Technology users have a variety of needs. Fully understanding users’ needs involves conducting user experience research (UXR), research “into all aspects of an end-user’s experience” of technology (Norman & Nielsen, 2017) and can be either quantitative or qualitative. For disabled technology users, barriers exist to accessibility of information more generally. Accessibility is defined as the “extent to which products, systems, services, environments, and facilities can be used by people from a population with the widest range of characteristics and capabilities to achieve a specified goal in a specified context of use” (ISO, 2023). Barriers for people with disabilities exist with respect to accessing accurate and useful health information (Estacio et al, 2019). For instance, blind people encounter barriers to web accessibility when searching health information online more generally (Choi et al, 2020), and deaf or hard of hearing folks encounter barriers when online content is not translated into American Sign Language (Kushalnagar et al, 2018). Intellectually disabled people face issues like lack of access to plain language summaries when seeking healthcare information as well (Dam et al, 2023). People with disabilities are therefore at a disadvantage when seeking health information because of a lack of societal consideration for their needs.

To provide a lens for evaluating current challenges that people with disabilities face while using technology to seek health information online, I bring UD into the conversation. Mace (1997) and Story and colleagues (1998) declare that UD is “the design of products and environments to be usable by all people, to the greatest extent possible, without the need for adaptation or specialized design.” I discussed the origins of UD in chapter 2. Today, UD also applies to digital environments (Rowland et al, 2010) and has been recommended in designing digital environments with respect to health information (Henni et al, 2022). Employing UD successfully in digital environments that health information can be accessed on requires input from people with disabilities (Henni et al, 2022). People with disabilities are the experts on themselves (Reaume, 2014) and their experiences must be centered when considering the barriers people encounter to accessing crucial health information (Kirschner et al, 2024). Otherwise, health information remains inaccessible to those most vulnerable. Inaccessibility of health information may contribute to delayed or no treatment for health problems especially in people with disabilities, resulting in severe illness or death (Kirschner et al, 2024).

It can therefore be inferred that the importance of accessibility of accurate health information in the age of AI cannot be underestimated when thinking of the user experience of people with disabilities who use technology. One related topic that intersects UD, UXR, AI, and health information is usability, which are discussed below.

### Usability and Its Relationship to Critical Disability Studies and Quantitative UXR

Usability is “the extent to which a product can be used by specified users to achieve specified goals with effectiveness, efficiency, and satisfaction in a specified context of use” (Grassi et al, 2017). Unusability, then, is its opposite. Different users experience the usability of a product to different degrees. As an example, screen readers are complex software that translates written text into voice text or Braille (Jordan et al, 2024). The two most widely used types of screen readers are JAWS and NVDA (Moncy et al, 2024). Someone who uses a JAWS screen reader might have a quite different experience of a given webpage than someone who uses an NVDA screen reader for the exact same webpage (Moncy et al, 2024). Thus, the reader can think of usability as a gradient that differs by person.

The field of disability studies uses wide range of theoretical approaches to analyze disability as a cultural, political, and social phenomenon, and to challenge the norms that keep people with disabilities disenfranchised (Hall, 2019). Incorporating universal design into technology is one way to challenge those norms. Jay Dolmage (2005) states that universal design is a framework that holds that disability must be welcome in society and accommodated for; he also states that usability itself stems from universal design. He goes on to say that allowing people with disabilities to address usability of digital content and other tools directly allows for more intentional co-creation of accessible digital content. In a later essay, Dolmage stated that multiple means of representation, expression, and engagement are crucial to applying universal design in a given context once the needs of the users are clear (2015). Allowing disabled voices into the conversation therefore welcomes their participation in society and in their learning processes; their participation is therefore part of the universal design framework.

UD also holds that people with disabilities should feel welcome to be research participants. Research on disabled user’s needs— UXR—can be either quantitative or qualitative. Scholarly critiques exist on the use of quantitative methods in disability research. Blanchard and colleagues (n.d.) argue that quantitative methods may have created disability as a specific social group that warrants studying and specific policies. They go on to state that quantification of disability in the 20th century both led to and justified the horrors of eugenics. They also state that the results and observations that were needed to make headway for disability protections to be established did not require quantitative methods. Additionally, the authors state that people who have endured discrimination in the past are harmed by bias in AI. Since AI is already linked to quantitative data (Torrentira, 2024), it makes sense that some healthy skepticism should exist with respect to quantitative research when it comes to disability.

Pamela Block and Gillian Parekh have some thoughts to offer, though, on the uses of quantitative research in and on disability, and so does A. Miller. Block states that while quantitative research may be considered as a compromising factor in disability studies research, such a mentality would suggest that one way of doing disability studies work is better than the other, when the reality is that there is no worse or better way to do disability research–in other words, “my disability studies is not better than yours” (2017). Parekh (2021) states that while there are ethical considerations to ensure that people with disabilities are not harmed in the process of research, especially if they are minors, conducting quantitative inquiry through a critical disability studies lens does indeed honor and center the experiences of people with disabilities (2023). A. Miller (2025)’s review paper highlights how to achieve inclusive survey instrument design by pulling from critical disability studies frameworks to ensure better user experience in people with disabilities who take surveys.

They also state that having inclusive survey design will enhance research participation in, and outcomes for, people with disabilities. Block, Parekh, and Miller therefore provide a useful guide as to how to conduct quantitative research in a way that honors disability studies and, as a result, the wishes of people with disabilities.

Since the goal for this article is to examine the user experience of people with disabilities encountering health information online about COVID-19 compared to their counterparts without disabilities, it is fair to consider what UXR methods would be appropriate to answer this question. Rohrer (2022) provides a guide as to when to use which UXR methods, stating that surveys are used to help track or discover critical issues to address. Additionally, Celentano and colleagues (2025) state that surveys will compare different segments of a given population at the same point in time. Since tracking and discovering important issues in accessibility, usability, and the online COVID-19 related content itself are the focus for this chapter, and we are comparing the experiences of people with and without disabilities, a survey is the best tool to use to this end.

### COVID-19 Information and Quantitative UXR

Since the announcement of the end of the public health emergency by the World Health Organization in 2023 (Alcendor et al, 2024), there has been scant information to assess if people with disabilities experience information on COVID-19 differently from people without them. COVID-19 remains a public health threat in both acute and long form, even if people do not understand or are not aware of the risks of the disease (Motta et al, 2025).

Furthermore, misinformation about the disease is still widespread (Kisa & Kisa, 2024). Assessing any disparities in health information receipt between people without and with disabilities about COVID-19 in the present day is therefore critical to maintaining not only the health of people with disabilities (Mills eds, 2025; Kuper & Smythe, 2023), but for overall population health (Kisa & Kisa, 2024).

People with disabilities are not a monolith in terms of accessibility challenges and needs related to health information. Visually impaired and blind people often struggle to use web-based tools (Sunkara et al, 2024) and health information more broadly (Choi et al, 2023). Good COVID-19 information, though applicable and critical to all people, often still is not accessible to people with disabilities (Jesus et al, 2023). Most older adults, who are more likely to have disabilities, are also not confident they could spot health misinformation from AI-generated tools (Keenan et al, 2024). Other discussions of challenges of people with disabilities accessing health information are presented in the sections above.

The above literature provides a general idea of issues people with disabilities face when trying to access information more generally or COVID-19 information specifically. But none of these studies discuss the usability of health information that people with disabilities, specifically, are accessing through the internet, often via embedded AI tools they may not even realize are being used, after the public health emergency ended.

Extant literature details the plight of the blind and visually impaired when it comes to accessing health information via technology (Sunkara et al, 2024; Moncy et al, 2024). The same extent of the body of literature does not necessarily exist for other disabilities. As a result of this literature gap, it is also unknown if usability impacts access to accurate information about COVID-19 transmission, symptoms, testing, treatment, and prevention—or, if usability could create differential access where some information is easier to access than other, possibly better quality, information about COVID-19.

Per the discussion above, experiences of people with disabilities when exposed to these AI-impacted messages through these mediums compared to the experiences of able-bodied people after the end of the public health emergency is unknown. Hence the research question below:

*To what extent do people with disabilities find accessing COVID-19 information to be both usable and satisfying when compared to people without disabilities?*

There are quantitative methods that can help answer the question above. One of those methods is a survey on user experience of health information on COVID-19. Both people with and without disabilities can take this survey so that the researchers can compare their experiences. This approach is discussed in more detail below.

### Data and Methods

Eligibility criteria for taking the survey were as follows: at least 18 years old, residents of the state of Virginia, able to understand written English, and had access to a computer, smartphone, tablet, or other device to take the survey. The study was declared IRB exempt (IRB #25-232). An even split between people with and without disabilities was targeted to allow for statistical comparison. Participants were recruited through student and disability-related listservs, on-campus fliers in college and/or university settings, and word of mouth.

Data was collected using a cross-sectional online survey from March to May of 2025 and administered via QuestionPro. Survey questions were drawn from the following sources: the US Census Bureau (2020), the System Usability Scale, or SUS (Hyzy et al, 2022), and the Health-ITUES (Schnall et al, 2018).

The SUS is a scale meant to measure how usable a given tool or system is for a user; a score of 68.0 out of 100 on the SUS is considered acceptably usable (Hyzy et al, 2022). The Health-ITUES is meant to measure how satisfying, in addition to usable, a given web-based or other technology is for viewing and managing health related content (Schnall et al, 2018). A score of 4.3 out of 5 is considered acceptably satisfying on the Health-ITUES (Loh et al, 2022). The full survey instrument can be found in Appendix 3A. We also adapted some questions from the American Community Survey (Altman et al, 2017) to ask participants about their disability status and type.

Respondents initially were asked several socio-demographic questions, including whether they are disabled and, if so, what categories their disabilities belong to. Categories of disability included blind/visually impaired, deaf/hard of hearing, mental health disability, developmental disability, and physical disabilities, among others; these categories were chosen based on the disability questions from the American Community Survey. Participants were also asked about their use of a variety of assistive tools and technology, ranging from screen readers and magnification to prosthetics and emotional support animals; these questions were developed by the research team.

Usability was measured with questions from the SUS. Further details can be seen in Appendix 3A. Traditional SUS scoring involves converting responses from a 5-point scale to adjusted points (0-4), summing them, and multiplying them by 2.5 to get a score out of 100. Traditional SUS scoring was used for this data.

Satisfaction with the online content that technology users found about COVID-19 was measured by the questions adapted from the Health-ITUES. Questions about content satisfaction were chosen to keep the focus of the participants on their satisfaction with COVID-19 related online content. The research team determined it was best to keep the ITUES scales out of 100 to keep pace with the SUS also being out of 100. The researchers therefore transformed the ITUES scale questions to match a 100-point scale, meaning a score of 86.0 was considered acceptably satisfying.

All survey takers were asked questions from the SUS and Health-ITUES for the internet’s use to access COVID-19 information and satisfaction with the content they found, respectively. Disabled survey takers who reported using at least one assistive technology were also asked questions from the SUS about their user experience of those assistive tools.

The survey remained open for about a month and took a mean of 5.93 minutes to complete. All statistical analysis was performed in R version 4.5.0 using RStudio software (Posit, 2025). Both the tidyverse (Wickham et al, 2019) and haven (Wickham et al, 2023) packages were also used in data analysis in conjunction with RStudio.

Criteria for inclusion in the statistical analysis included the following: consented to the study participation, reached the last question asked of all participants, and used online sources to find health information on COVID-19 at least once after the public health emergency declared by WHO ended.

## Results

A total of 172 participants met the inclusion criteria. Table 1 shows the socio-demographic breakdown of those 172 participants.

**Table 1:** Socio-Demographic Characteristics of Respondents (N=172) Note: Percentages may not add to 100.

|  | Frequency | Percent (%) |
| --- | --- | --- |
| *Age* |  |  |
| 18-24 | 41 | 23.8 |
| 25-34 | 48 | 27.9 |
| 35-44 | 22 | 12.8 |
| 45 and up | 61 | 35.5 |
| *Race/Ethnicity* |  |  |
| Asian or Middle Eastern | 14 | 8.1 |
| Caucasian or White (non-Hispanic) | 142 | 82.6 |
| Black non-Hispanic | 5 | 2.9 |
| Hispanic/Latinx | 6 | 3.5 |
| Native Hawaiian/Pacific Islander and American Indian | 0 | 0 |
| Multiracial | 4 | 2.3 |
| Prefer not to say | 1 | 0.6 |
| *Gender* |  |  |
| Man | 31 | 18.0 |
| Woman | 129 | 75.0 |
| Transgender or other | 12 | 7.0 |
| *Education Level* |  |  |
| Less than high school | 0 | 0 |
| HS or equivalent and 2-year college | 14 | 8.1 |
| Some college | 21 | 12.2 |
| 4-year college | 47 | 27.3 |
| Some graduate school | 19 | 11.0 |
| Graduate or professional degree | 71 | 41.3 |
| *Disability Status* |  |  |
| Not disabled | 96 | 55.8 |
| Disabled | 76 | 44.2 |
| *Disability Type* |  |  |
| *Note: some people have multiple disabilities.* |  |  |
| Hearing | 5 | 2.9 |
| Visual | 3 | 1.7 |
| Developmental | 39 | 22.7 |
| Communication | 3 | 1.7 |
| Chronic Illness | 19 | 11.0 |
| Learning | 7 | 4.1 |
| Intellectual | 0 | 0 |
| Mental Health | 35 | 20.3 |
| Mobility | 5 | 2.9 |
| Neurological | 6 | 3.5 |
| Other | 3 | 1.7 |
| *Accommodations Reported in Disabled Participants* |  |  |
| *Note: may total to more than 100%\*\** |  |  |
| Screen magnification or large print | 11 | 14.5 |
| Braille | 1 | 1.3 |
| Communication Access in Real Time (CART) | 1 | 1.3 |
| Screen reader | 3 | 3.9 |
| Closed captions | 33 | 43.4 |
| Speech to text | 6 | 7.9 |
| Webpage tools | 3 | 3.9 |
| Dyslexia friendly interface | 1 | 1.3 |
| Sign language or sonification | 0 | 0 |
| Cane, Crutch, or Wheelchair | 7 | 9.2 |
| Service Animal or Prosthetic | 0 | 0 |
| Emotional Support Animal | 6 | 7.9 |
| Other | 6 | 7.9 |
| None | 28 | 36.8 |

\*\*Some people reported the use of more than one of the listed accommodations.

For comparison’s sake, the United States Census Bureau (2020) reported the state of Virginia has a population of 8,631,393 people, and that 42.4% of Virginians have a bachelor’s degree or more; 908,749 Virginians report being Hispanic/Latino; and the median age of a Virginian is 39.3 years. Per the US census for the entire country (2020), the US median age is 39.2 years; 36.2% of people have a bachelor’s degree or more, and the total population is 331,449,281 people; 62,080,044 people reported being Hispanic/Latino for the entire US census of 2020. This sample is comparable to the age and educational attainment of the rest of the state as of 2020, but not the rest of the US as of 2020, especially when it comes to education level. Fewer people reported being Hispanic/Latino in this sample than in either the state of Virginia or the US overall, which also means viewing these results with caution when it comes to racial and ethnic makeup.

Of the 172 participants noted above, 142 answered the usability and satisfaction scoring questions detailed in Summaries 2A and 2B.

### Summary 2A: Descriptives and Test for Difference in Usability Scores between People with and Without Disabilities (N=142)

Summary 2A details the results of the usability scores derived from the questions pulled from the SUS for both disabled and nondisabled users, with a 68.0 score being considered acceptably usable. The median usability score for this study was 71.25; 25^th^ percentile score was 62.5, and 75^th^ percentile score was 80, with a minimum score of 32.5 and a maximum score of 100.

Among those without disabilities, 52 out of 74 (70.3%) found both internet-based tools and information about COVID-19 usable. By contrast, only 52.9% of people with disabilities found both internet-based tools and information about COVID-19 usable.

Comparisons were also made between participants with and without disabilities in this study via Mann-Whitney tests. The Mann-Whitney tests were run as an alternative to t-tests at the 0.05 level after the Shapiro test showed that SUS scores were not normally distributed and the test for equal variances showed that they were not equal.

*Mann-Whitney Test for Usability Scores between People with and without Disabilities*

W = 2125.5, p-value = 0.1102

There is not a statistically significant difference in the mean usability scores of tools and sources to access COVID-19 information between people with and without disabilities.

Summary 2B: Test for Satisfaction with Information Scores between People with and Without Disabilities (N=142)

Summary 2B details the results of the satisfaction scores derived from the questions adapted from the Health-ITUES for both disabled and non-disabled users, with a score of 86.4 being considered acceptable. The median score was 60; 25^th^ percentile score was 55; 75^th^ percentile was 80, with the minimum score being 20 and the maximum being 100.

Again, Mann-Whitney tests were run as an alternative to t-tests at the 0.05 level after the Shapiro test showed that SUS scores were not normally distributed and the test for equal variances showed that they were not equal.

*Mann-Whitney for Satisfaction Scores between People With and Without Disabilities:*

*W = 1026.5, p-value = 0.01047*

There is a statistically significant difference in the mean satisfaction scores with respect to accessing COVID-19 information between people with and without disabilities.

Due to limitations in sample size and power, significance was assessed using bivariate correlations. Table 2 is a table of bivariate correlations that shows the strength of association and linear relationship between variables, as seen below. All significance is reported at the 0.05 level per statistical convention.

**Table 2:**
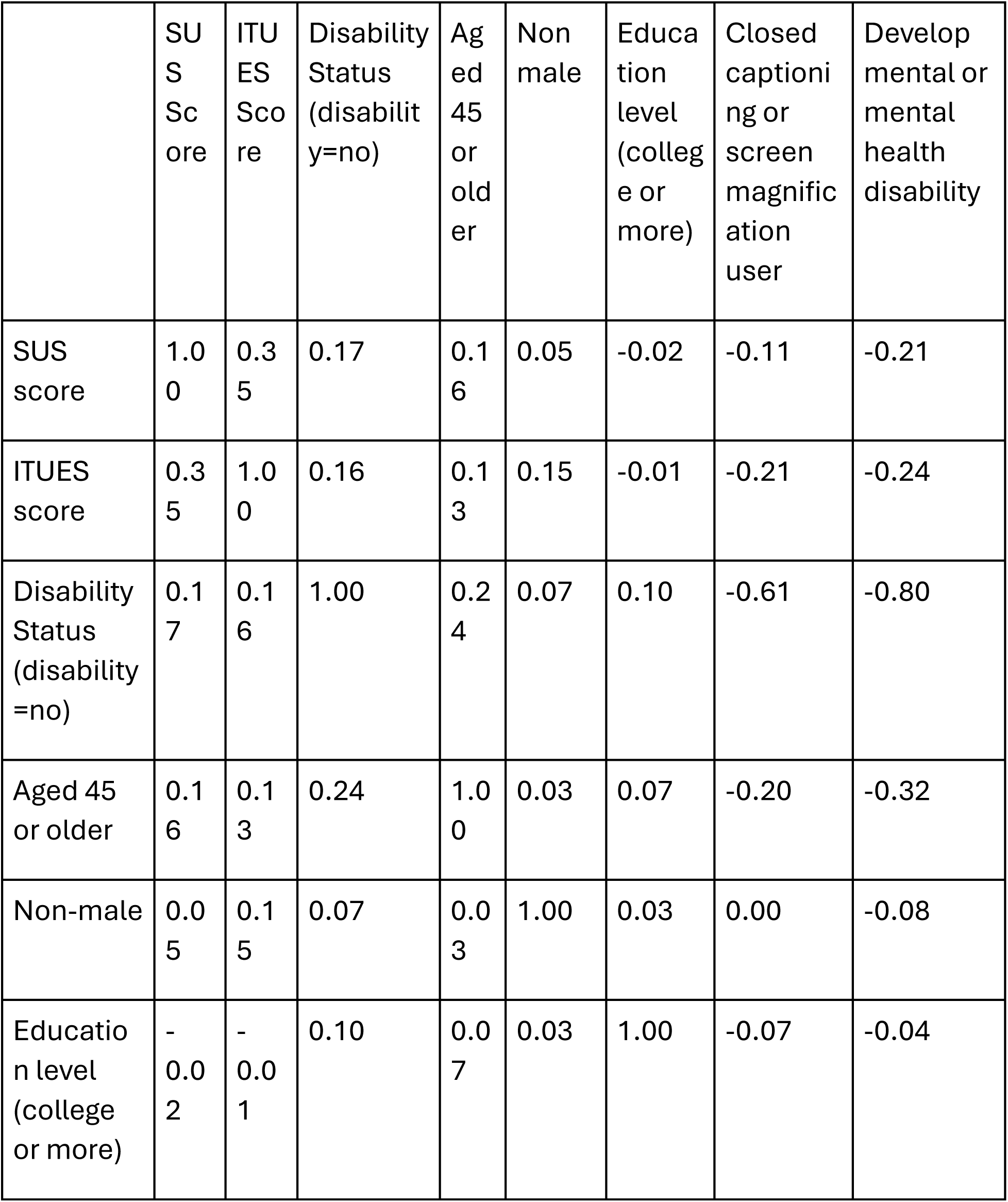
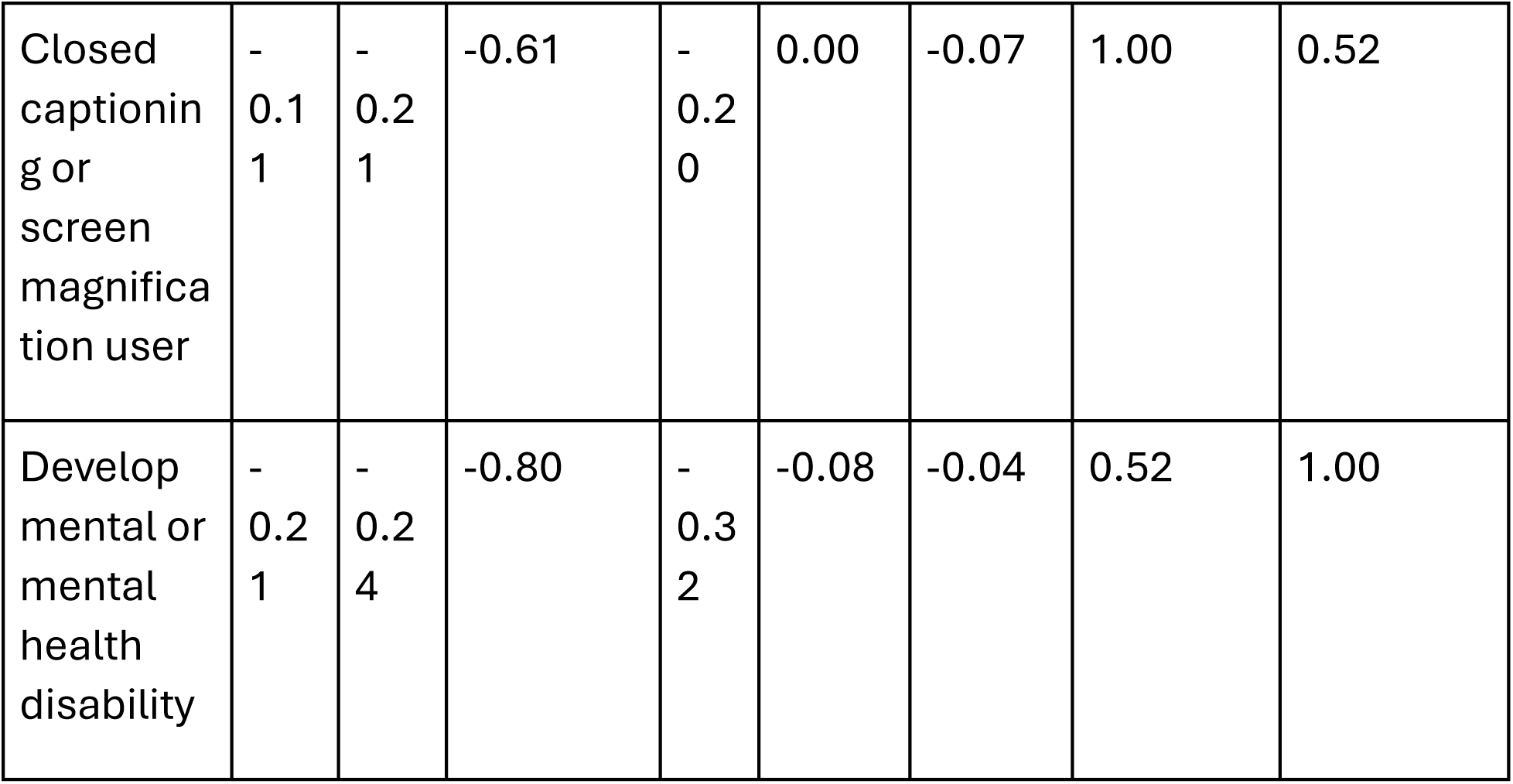
Bivariate Correlations among Variables (N=142).

|  | SUS<br>Score | ITU<br>ES<br>Score | Disability<br>Status<br>(disability=no) | Aged<br>45<br>or<br>older | Non<br>male | Educational<br>level<br>(college or<br>more) | Closed<br>captioning<br>or<br>screen<br>magnification<br>user | Developmental<br>or<br>mental<br>health<br>disability |
| --- | --- | --- | --- | --- | --- | --- | --- | --- |
| SUS<br>score | 1.00 | 0.35 | 0.17 | 0.16 | 0.05 | -0.02 | -0.11 | -0.21 |
| ITU<br>ES<br>score | 0.35 | 1.00 | 0.16 | 0.13 | 0.15 | -0.01 | -0.21 | -0.24 |
| Disability<br>Status<br>(disability=no) | 0.17 | 0.16 | 1.00 | 0.24 | 0.07 | 0.10 | -0.61 | -0.80 |
| Aged 45<br>or older | 0.16 | 0.13 | 0.24 | 1.00 | 0.03 | 0.07 | -0.20 | -0.32 |
| Non-male | 0.05 | 0.15 | 0.07 | 0.03 | 1.00 | 0.03 | 0.00 | -0.08 |
| Educational<br>level<br>(college<br>or more) | -0.02 | -0.01 | 0.10 | 0.07 | 0.03 | 1.00 | -0.07 | -0.04 |
| Closed captioning or screen magnification user | -0.11 | -0.21 | -0.61 | -0.20 | 0.00 | -0.07 | 1.00 | 0.52 |
| Developmental or mental health disability | -0.21 | -0.24 | -0.80 | -0.32 | -0.08 | -0.04 | 0.52 | 1.00 |

Usability scores were significantly lower in people who had developmental or mental health disabilities (r=-0.21, p=0.0131). Satisfaction scores were significantly lower in people who had a developmental or mental health disability (r=-0.24, p=0.0111) and in those who used screen magnification or closed captioning (r=-0.21, p=0.0262).

Ordinary least squares (OLS) regression results pertaining to both usability of tools to access COVID-19 information (from the SUS) and satisfaction with COVID-19 information itself (from the ITUES) are shown in table 3.

**Table 3:** OLS Regressions of System Usability Score (SUS) of COVID-19 Information and Satisfaction with COVID-19 Information (N=142).

| Model 1: OLS Regression for SUS of COVID-19 Information |  | Model 2: OLS Regression for Satisfaction with COVID-19 Information |  |
| --- | --- | --- | --- |
| Variable | Coefficient | Variable | Coefficient |
| *Age (ref=<45)* |  | *Age (ref=<45)* |  |
| 45 and up | 3.04 | 45 and up | 1.385 |
| *Gender (ref=male)* |  | *Gender (ref=male)* |  |
| Non-male (includes female, transgender/other) | 0.59 | Non-male (includes female, transgender/other) | 5.23 |
| *Education Level (ref=some college or less)* |  | *Education Level (ref=some college or less)* |  |
| College or more | -0.74 | College or more | -1.64 |
| *Disability Status (ref=no disability)* |  | *Disability Status (ref=no disability)* |  |
| Disabled | 0.83 | Disabled | -5.38 |
| *If disabled, type of dis (ref=something else)* |  | *If disabled, type of dis (ref=something else)* |  |
| Developmental or mental health | -4.29 | Developmental or mental health | -8.134 |

| *If indicated, Type of Accommodation (ref=something else)* |  | *If accommodation, type (ref=something else)* |  |
| --- | --- | --- | --- |
| Closed captioning or screen magnification | 0.02 | CC or Screen Mag | -6.80 |
| *Constant* | 71.43 | Constant | 65.31 |
| AIC | 1159 | AIC | 889.9 |

Table 3 should be interpreted cautiously, since the sample did not have sufficient power to produce any significant regression coefficients. Power and sample size issues are common issues in user study research; smaller sample sizes are acceptable for formative UX research (Budiu & Moran, 2021). Note that statistical significance was set to 0.05 for table 3.

For model 1, degrees of freedom=141 total, 135 residual. For model 2, degrees of freedom=108 total, 102 residual.

The model predicts that a non-disabled, non-college educated, white, able-bodied male who used no accommodations on average has an SUS score of 71.43 out of 100, where the scale is designed such that a score of 68.0 is considered acceptably usable. They would also have a Health-ITUES score of 65.31 out of 100 when the scale is designed such that a score of 86.0 out of 100 is acceptably satisfying.

On average, model 1 predicts that those who reported having developmental or mental health disabilities would have a 4.29-point lower usability score than people without developmental or mental health disabilities, and those who reported having a college degree or more would likely report a 0.74-point lower usability score than those who reported having less education.

On average, model 2 (for satisfaction score) predicts that those with a college degree are likely to report a 1.64-point lower satisfaction score compared to those with less than a college degree. People with disabilities are likely to report a 5.38-point lower satisfaction score compared to those who reported no disabilities. People who use closed captioning or screen magnifiers/large print are likely to report 6.8 points less satisfaction with online content than those who did not report using either assistive tool. People who have developmental or mental health disabilities are likely to report an 8.134-point lower satisfaction score than those who do not have developmental or mental health disabilities.

Both models indicate that while many people find tools to help information usability, the degree to which they find the content they encounter satisfying varies widely between individuals and groups, including based on type of disability and the tools being used. Both models also indicate that people with disabilities have more difficulty using technology, and finding content usable and satisfying, compared to their counterparts that do not have disabilities.

## Discussion

The results of the quantitative study presented above indicate that participants with disabilities found technology more difficult to use–and content less satisfying—than participants without them. The most frequently reported disabilities were developmental and mental health disabilities. The most frequently reported accommodations in people with disabilities that participated in this study were closed captioning, screen magnification, and large print. Most participants, regardless of disability status, are dissatisfied with COVID-19 information found on social media.

This study should be treated as a pilot study and regarded with caution. In addition to its limited sample size and less than representative sample, other factors should be considered when considering the results. For one thing, these participants had access to the internet and/or AI influenced tools. People with disabilities are less likely to report access to these tools than people without disabilities (Perrin & Atske, 2021). Additionally, many of the survey participants did not identify as black, Hispanic, or indigenous–or any nonwhite group for that matter. Also, most participants reported college degrees; as of 2022, only 23.5% of US adults had a bachelor’s degree (US Census Bureau, 2022) despite scholarly and public knowledge that education affords more access to systems and opportunities (Mitra, 2011). It makes sense that results would look different in a sample which reports less formal education and/or more racial diversity. Further research should aim to recruit a more racially and educationally diverse sample and highlight where the differences fall.

One unexpected finding that could benefit from follow-up research is the drop in both general usability and satisfaction with the content received in people with more education. People with higher education levels may be using more complex software than accessibility tools, at present, can keep up with.

People with a college degree may also have higher expectations of the level of usability they should find. Because they have more resources and more training and exposure to what could be, they may have higher standards.

Other limitations exist in this study. For ease of data collection, the study was limited to Virginia residents, so systematic differences between Virginia (or this sample of Virginians) and the US population limit its generalizability. Also, current data does not allow examination of the connection between satisfaction with the content received and the real or perceived value of that content to the user. Nonetheless, people with disabilities are more likely to be dissatisfied with the content they receive about COVID-19, compared to their peers without disabilities. They also encounter general usability issues with technology even with accommodations in place. Reasons for these usability and satisfaction issues vary and require follow-up research conducted qualitatively with those technology users.

### Conclusions

Both usability and content are important aspects of technology use for health information seeking, both generally and in relation to COVID-19. This survey provides a starting point to estimate both usability and satisfaction of content with respect to COVID-19 information and suggest key categories of heterogeneity to consider when studying health information usability in both research and applied settings.

Participants overall reported being dissatisfied with the content itself about COVID-19, especially on social media. However, participants with disabilities were more likely to report both issues with both usability and satisfaction with COVID-19 content they found online compared to their counterparts without disabilities, even with their accommodations in place. These results establish the difference between people with disabilities who aim to access high quality health information online in contrast to their peers without disabilities. In other words, people with disabilities continue to be left at a disadvantage compared to people without disabilities when using technology to access content regarding COVID-19 online. Further work should involve recruitment of people with and without disabilities in a more racially and educationally diverse context for further insights to see if similar results would be found.

Further work should also involve qualitative research with technology users to determine specific reasons for user dissatisfaction.

## Data Availability

All data produced in the present study are available upon reasonable request to the authors.

